# Four-State Hemodynamic Framework for Neovascular Age-Related Macular Degeneration: Mathematical Model Integrating Vascular Permeability, Lesion Transmural Pressure, Choroidal Excess Pressure, and Wall Stability

**DOI:** 10.64898/2026.10.05.26364815

**Authors:** Yasuo Yanagi, Satoru Inoda, Keiko Azuma, Hidenori Takahashi, Tatsuya Inoue, Maiko Maruyama-Inoue, Kazuaki Kadonosono

## Abstract

**Purpose:** To apply a mathematical model integrating vascular permeability, lesion transmural pressure, choroidal excess pressure, and wall stability to incomplete fluid response and massive submacular hemorrhage in neovascular age-related macular degeneration.

**Methods:** Permeability was calculated from aqueous vascular endothelial growth factor (VEGF). Windkessel gave usual lesion pressure from choriocapillaris pressure and high inflow from an arterial-level puncture pressure used as a working value. Choroidal excess pressure was calculated from choroidal thickness and a pachy index as increased outflow resistance. Exudative change used a revised Starling equation scaled to early central-retinal-thickness reduction, with 40-μm threshold. A relative rupture-hazard index was calculated from Laplace wall stress and wall stability. VEGF, pressures, and that reduction were from the literature. Pressure was isolated from permeability and choroidal excess.

**Results:** In the model, raising lesion pressure from choroidal-excess lesion pressure to high inflow increased untreated exudative change from 150 to 760 μm. Residual under saturation was 64 μm as a lower-end reference (Monte Carlo 69-286 μm; median, 170) and 90 μm with high-hazard choroidal excess; both above 40 μm. Reducing choroidal thickness in the thickened choroidal-excess state added 13-14 μm, smaller than that residual. Lowering lesion pressure from high inflow to usual lesion pressure (21 to 10 mmHg) reduced the index to 0.64 of the high-hazard reference, and to 0.34 with wall stabilization.

**Conclusion:** Lesion transmural pressure due to high inflow left residual exudation even after permeability suppression with anti-VEGF. Fluid resolution on optical coherence tomography may not parallel a reduction in the rupture-hazard index, depending on wall stability.

## Introduction

Neovascular age-related macular degeneration (nAMD) is commonly classified by the anatomical location of macular neovascularization (MNV), including type 1, type 2, and type 3 MNV as described by Freund and colleagues. Polypoidal choroidal vasculopathy (PCV) and pachychoroid neovasculopathy (PNV) are generally considered in parallel with this framework [1–4]. Treatment is based principally on suppression of vascular endothelial growth factor (VEGF), with some agents also targeting angiopoietin-2 (Ang2). Comparisons among agents therefore often emphasize binding affinity, pharmacokinetics, and the speed or durability of fluid resolution [5][6].

In this regard, two points require attention.

First, there is little theoretical account of how hemodynamics might explain incomplete treatment response. Direct measurements of effective driving pressure in MNV are not available, yet a Starling-type formulation requires permeability and that driving pressure [7][8]. In the absence of such measurements, we assumed that effective driving pressure depends on both lesion transmural pressure, set by inflow relative to outflow, and choroidal excess pressure from increased outflow resistance with choroidal thickening [8][9]. There is great variability in choroidal thickness in patients with MNV [4][10] and, possibly, in the pressure feeding the lesion [11][12]. Usual lesion pressure is at the choriocapillaris level; as suggested by histologic studies of intact eyes, the lesion may instead connect to a posterior ciliary arteriole or artery [13][14].

Second, VEGF concentration and binding affinity alone may not account for hemorrhage, because rupture depends on wall stress relative to wall strength, as a thin-wall Laplace model predicts [15][16]. Serous exudation and vascular rupture are related but nonidentical physical processes, and a reduction in optical coherence tomography (OCT) fluid need not imply a proportional reduction in SMH risk. In support of this, the 10-year cumulative incidence of massive SMH was higher in PCV than in typical nAMD (29.9% vs 9.9%; hazard ratio, 2.66) [17], whereas aqueous VEGF in eyes with massive hemorrhage was low (median, 5.2 pg/mL) [18].

On this basis, we developed a four-state hemodynamic framework that integrates vascular permeability, lesion transmural pressure, choroidal excess pressure, and wall stability: high-hazard, residual-exudation, choroidal-excess, and low-choroidal-excess (Table 1 and Figure 1), intended to complement anatomical MNV typing. The primary outputs were foveal exudative change and a relative rupture-hazard index. The model tested three questions: (1) the effect of raising lesion pressure to the highest value on exudative change; (2) the incremental effect of reducing choroidal thickness on exudative change in the choroidal-excess and low-choroidal-excess states; and (3) the effect of reducing lesion pressure with or without wall stabilization on the relative rupture-hazard index. It predicts that persistent exudation after marked permeability suppression with anti-VEGF treatment may reflect a pressure-related component, and that resolution of exudation need not indicate a reduction in rupture propensity.

**Figure 1.**
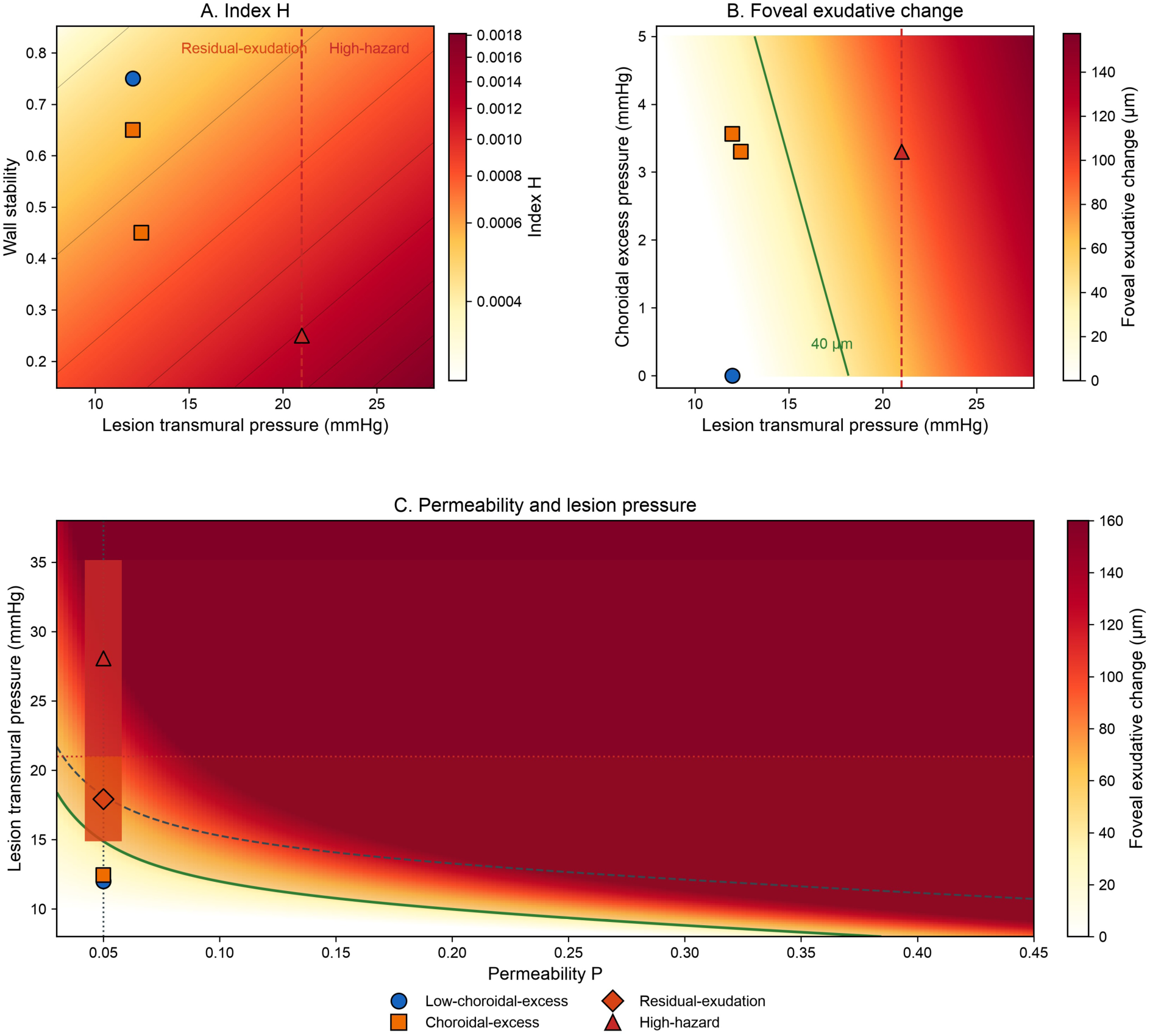
Classification of hemodynamic states. A: The index H in Methods 6 as a continuous quantity of ΔPlesion (horizontal axis) and wall stability (vertical axis). Color is H, darker at higher index. The orange band is residual-exudation (about 15 mmHg to below 21 mmHg); the red band and dashed line are the high-hazard state from 21 mmHg. B: Foveal exudative change under pharmacological saturation (D=10^6^ pM) as a continuous quantity of ΔPlesion and choroidal excess pressure. The green contour is 40 μm. C: Foveal exudative change in the plane of permeability P and ΔPlesion. The vertical dotted line is the saturation permeability floor (Pbase). The solid and dashed green lines are 40 μm with the excess of the high-hazard row and with that excess set to zero, respectively. The orange band is the residual-exudation interval from the 40-μm boundary to less than 21 mmHg, drawn with the output of the high-hazard row. The red band is the high-hazard interval from 21 mmHg to the high-hazard upper bound.

**Table 1.**
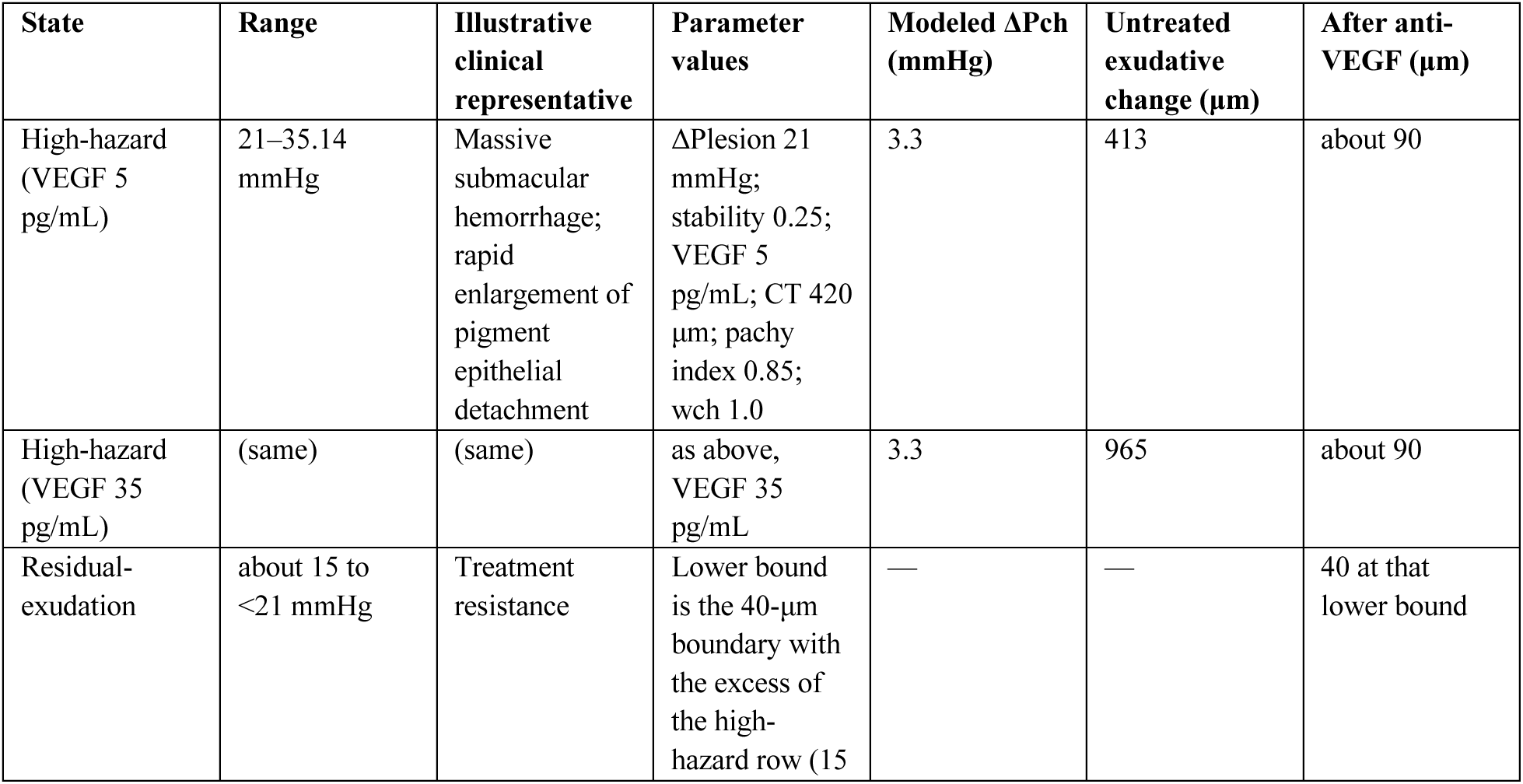
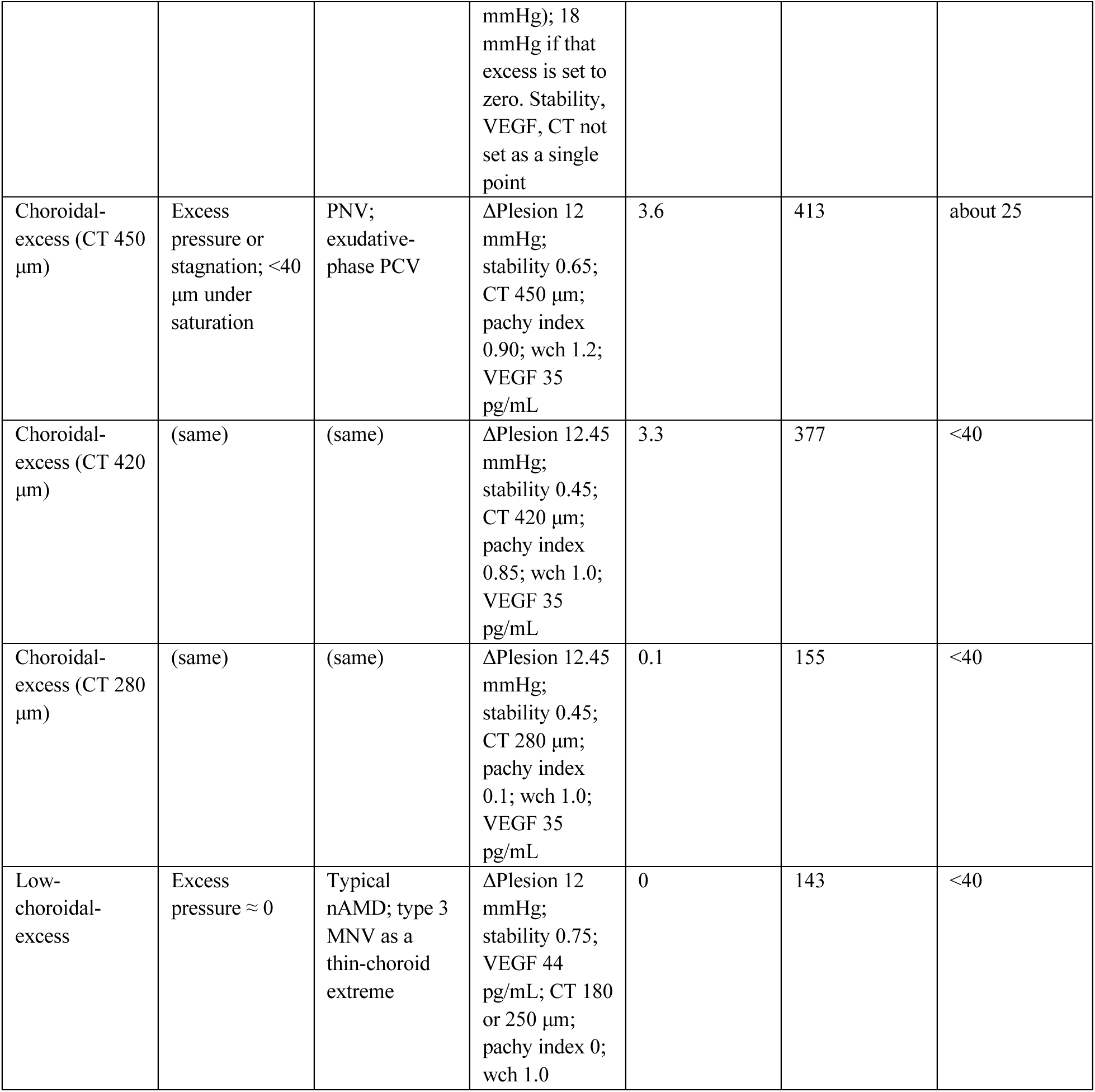
Hemodynamic states, parameter values, and model outputs.

| State | Range | Illustrative clinical representative | Parameter values | Modeled $\Delta P_{ch}$ (mmHg) | Untreated exudative change ($\mu m$) | After anti-VEGF ($\mu m$) |
| --- | --- | --- | --- | --- | --- | --- |
| High-hazard (VEGF 5 pg/mL) | 21–35.14 mmHg | Massive submacular hemorrhage; rapid enlargement of pigment epithelial detachment | $\Delta P_{lesion}$ 21 mmHg; stability 0.25; VEGF 5 pg/mL; CT 420 $\mu m$; pachy index 0.85; wch 1.0 | 3.3 | 413 | about 90 |
| High-hazard (VEGF 35 pg/mL) | (same) | (same) | as above, VEGF 35 pg/mL | 3.3 | 965 | about 90 |
| Residual-exudation | about 15 to <21 mmHg | Treatment resistance | Lower bound is the 40-$\mu m$ boundary with the excess of the high-hazard row (15 | — | — | 40 at that lower bound |
|  |  |  | mmHg); 18 mmHg if that excess is set to zero. Stability, VEGF, CT not set as a single point |  |  |  |
| Choroidal-excess (CT 450 $\mu\text{m}$) | Excess pressure or stagnation; <40 $\mu\text{m}$ under saturation | PNV; exudative-phase PCV | $\Delta\text{Plesion}$ 12 mmHg; stability 0.65; CT 450 $\mu\text{m}$; pachy index 0.90; wch 1.2; VEGF 35 pg/mL | 3.6 | 413 | about 25 |
| Choroidal-excess (CT 420 $\mu\text{m}$) | (same) | (same) | $\Delta\text{Plesion}$ 12.45 mmHg; stability 0.45; CT 420 $\mu\text{m}$; pachy index 0.85; wch 1.0; VEGF 35 pg/mL | 3.3 | 377 | <40 |
| Choroidal-excess (CT 280 $\mu\text{m}$) | (same) | (same) | $\Delta\text{Plesion}$ 12.45 mmHg; stability 0.45; CT 280 $\mu\text{m}$; pachy index 0.1; wch 1.0; VEGF 35 pg/mL | 0.1 | 155 | <40 |
| Low-choroidal-excess | Excess pressure $\approx 0$ | Typical nAMD; type 3 MNV as a thin-choroid extreme | $\Delta\text{Plesion}$ 12 mmHg; stability 0.75; VEGF 44 pg/mL; CT 180 or 250 $\mu\text{m}$; pachy index 0; wch 1.0 | 0 | 143 | <40 |

## Methods

### 1. Study Design

This was a theoretical modeling study. The three questions are stated in the Introduction. Computational implementation, extraction of numerical outputs, and generation of figures were performed with assistance from Cursor software agents (Cursor, version 3.20.17; Anysphere, Inc., San Francisco, CA, USA). All equations, inputs, outputs, and figures were reviewed and verified by the authors. Unless identified as from the literature, numerical inputs were chosen for the model.

### 2. End Points

Modeled foveal exudative change was calculated from the steady-state filtration volume of the revised Starling equation, converted to a thickness-equivalent value in micrometers, and scaled to the early reduction in central retinal thickness reported in CATT, a typical-nAMD trial [19][20]. Resolution of exudation was defined as a value below a 40-μm threshold and was varied from 30 to 60 μm in sensitivity analyses. The residual-exudation lower pressure boundary was the lesion pressure yielding residual exudation at that threshold under pharmacological saturation; this boundary was calculated with the choroidal excess of the high-hazard state and again after that excess was set to zero. The relative rupture-hazard index was calculated from Laplace wall stress and wall stability. Analyses used ratios of this index.

### 3. Hemodynamic States

Numerical inputs are in Supplementary Table S1. Choroidal excess pressure is defined in Methods 5-2. The high-hazard upper bound is the steady lesion pressure when raised inflow-side and outflow-side resistance (stagnation) are superimposed on high inflow. For the representative high-hazard analysis, lesion pressure was fixed at high inflow and aqueous VEGF was varied.

### 4. Vascular Permeability Index

We derived a permeability index P (0 to 1) from a systems-biology model of angiogenic signaling [21] that included VEGF-dependent Src activation, Ang1–Tie2 stabilization, and Ang2 antagonism of Tie2. The signaling equations are in Supplementary Table S2. For high drug concentrations, the free-VEGF fraction was

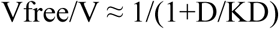

where D and KD were expressed in pM. Pharmacological saturation used aflibercept at D=10⁶ pM [5]. P served as the filtration coefficient [7].

### 5. Lesion Transmural Pressure and Modeled Exudative Volume

Lesion transmural pressure was calculated from the steady intravascular lesion pressure

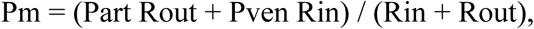

where Part is inflow pressure, Pven is venous pressure, and Rin and Rout are the inflow-side and outflow-side resistances. Transmural pressure was ΔP=Pm-Ptissue. Tissue pressure was set so that, with arteriolar-side and venous pressures from the literature, calculated steady transmural pressure matched the reported mean choroidal transmural pressure [9].

### 5-1. Modeled Inflow

Usual lesion pressure was calculated from arteriolar-side choriocapillaris pressure, and high inflow from an arterial-level puncture pressure used as a working value, both in the literature [9]. That working value corresponds to the posterior-ciliary connection in the Introduction [13][14]. Lesion pressure in the choroidal-excess state used the PLANET week-52 order of central-subfield-thickness reduction for the higher value [22].

A pressure reduction from high inflow to usual lesion pressure represented polyp closure.

### 5-2. Modeled Outflow and Choroidal Excess Pressure

Venous pressure used the intraocular-pressure reference plus a rabbit choroidal venous increment [8][9]. Outflow impairment raised outflow-side resistance. For stagnation, inflow-side resistance was the sum of feed and lesion-margin resistances; outflow-side resistance was drainage multiplied by a dimensionless tortuosity factor [23][24].

We calculated choroidal excess pressure (ΔPch) from choroidal thickness (CT, μm) and a pachy index p (0 to 1). Outflow resistance was scaled by

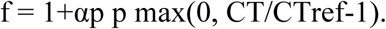

ΔPch was the difference between transmural pressure at this f and at f=1. Effective driving pressure for filtration was ΔPeff=ΔPlesion+wchΔPch. State CT and pachy-index values were chosen for the model, except that the thin-choroid low-choroidal-excess state used a choroidal-thickness value from the literature [10]. To represent a reduction in choroidal thickness after anti-VEGF treatment, CT and the pachy index were multiplied by published ratios for subfoveal choroidal thickness and choroidal vascularity index [26]. We calculated the incremental change in exudative change with permeability held constant.

### 5-3. Modeled Exudative Volume

The Starling model gives the steady-state filtration volume as

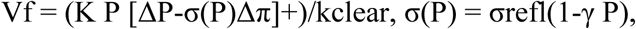

where [x]+=max(x,0). Modeled foveal exudative change was Vf· c, with c calibrated so that filtration in the low-choroidal-excess state matched the early reduction in central retinal thickness reported in CATT [19][20].

### 6. Wall stress and Relative Rupture-Hazard Index

Wall stress used a thin-wall Laplace form, σwall=ΔP r/(2hwall). Maximum wall stress was σmax=σstr(0.2+0.8 W) with W between 0 and 1. The index was

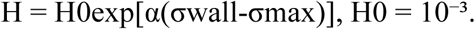

The index depends on wall stress and wall stability. When wall stabilization was added to the pressure reduction from high inflow, wall stability was set to that of the low-choroidal-excess state. Wall stress was related to wall strength following the peak-wall-stress and peak-wall-rupture-index literature for abdominal aortic aneurysm [15][16]. The coefficient α was calculated from the published hazard ratio of 2.66 for massive SMH between PCV and typical nAMD [17] and the stress difference between the high-hazard and low-choroidal-excess states, onto which those clinical groups were mapped. Subsequent values were interpolations within this calibrated exponential.

### 7. Sensitivity Analyses

Parameters were varied one at a time across predefined ranges, and 4000 sets were drawn from uniform distributions over the same ranges. Predefined outputs were the 40-μm boundary pressure, residual exudative change at high inflow with choroidal excess set to zero and pharmacological saturation, and the relative rupture-hazard index after the pressure reduction, with and without wall stabilization. Sampling ranges and replacement of the high-inflow working value are in the extended sensitivity methods and Supplementary Tables S4 and S5.

## Results

### 1. Lesion Pressure and Modeled Exudative Change

In the model, lesion pressure was approximately 10 mmHg under the usual condition, 13 mmHg with outflow impairment, and 21 mmHg at high inflow (Figure 2A). Effective transmural pressure over the cardiac cycle is shown in Video, Supplemental Digital Content 1. Parameter values and outputs for each state are given in Table 1.

**Figure 2.**
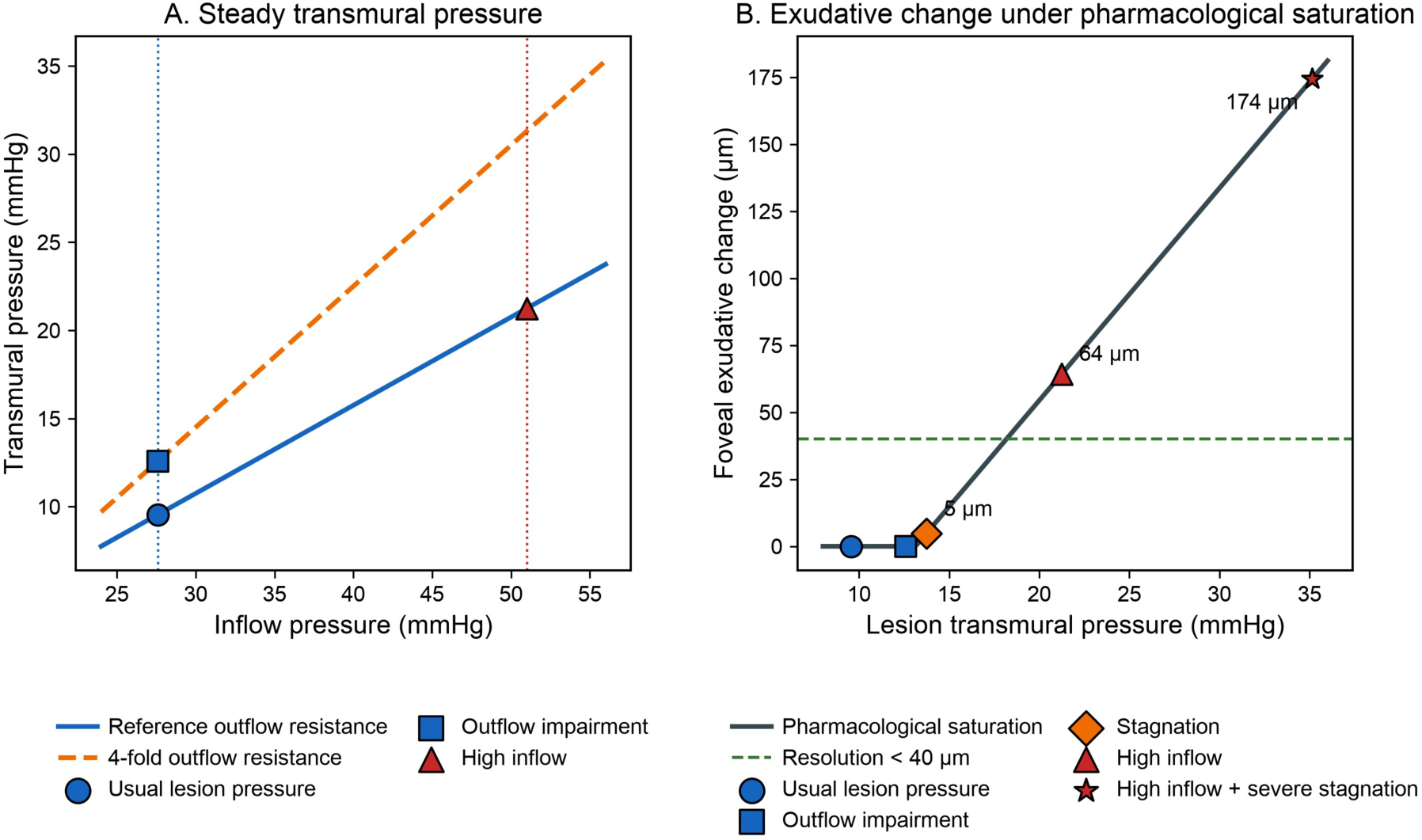
Steady transmural pressure against inflow pressure at two outflow resistances, and exudative change under pharmacological saturation. A: Steady transmural pressure versus inflow pressure. The solid line is reference outflow resistance; the dashed line is 4-fold outflow resistance. The vertical dotted lines are the usual arteriolar-side inflow and the high-inflow working value. B: Foveal exudative change versus ΔPlesion under saturation with choroidal excess set to zero. Residual values exceed 40 μm under high inflow (about 64μm) and at the high-hazard upper bound (about 174 μm at 35 mmHg). Stagnation alone is about 5 μm at 14 mmHg.

Pressure was isolated from permeability and choroidal excess. When permeability was held at the choroidal-excess value and choroidal excess pressure was set to zero, raising lesion pressure from the choroidal-excess lesion pressure to high inflow increased untreated foveal exudative change from approximately 150 to 760 μm. That isolation was repeated at the lower permeability of the high-hazard state. With permeability fixed at the high-hazard value corresponding to VEGF of 5 pg/mL, the same pressure pair raised the value from approximately 17 to 311 μm. Those isolation values are marked in Results 3.

When the combined values in Table 1 are applied for each state, the lesion pressure at which residual exudation remained at the resolution threshold under saturation was approximately 15 mmHg if the choroidal excess of the high-hazard state was included, and approximately 18 mmHg if that excess was set to zero (Figure 1C). Figure 1B is the same saturation field against lesion pressure and choroidal excess pressure. Residual thickness under saturation was then reported for each condition. Residual exudative change was below that threshold under the reference, outflow-impairment, and choroidal-excess conditions (approximately 5 μm with stagnation alone at 14 mmHg). It remained above the threshold at high inflow (approximately 64 μm with choroidal excess set to zero; approximately 90 μm at the high-hazard output in Table 1) and at the high-hazard upper bound (approximately 174 μm at 35 mmHg; Figure 2B). After permeability was suppressed, residual fluid persisted only when lesion pressure was raised to high inflow.

### 2. Incremental Effect of Reduced Choroidal Thickness

In the low-choroidal-excess state, ΔPch was 0 mmHg at CTs of 180 and 250 μm. Under these saturated conditions, the exudative change was below the resolution threshold, and further CT reduction produced no additional decrease (Figure 3A). The same CT reduction was then applied in a near-zero-excess condition with a CT of 280 μm. In this condition, ΔPch was 0.1 mmHg, and the incremental decrease was negligible.

**Figure 3.**
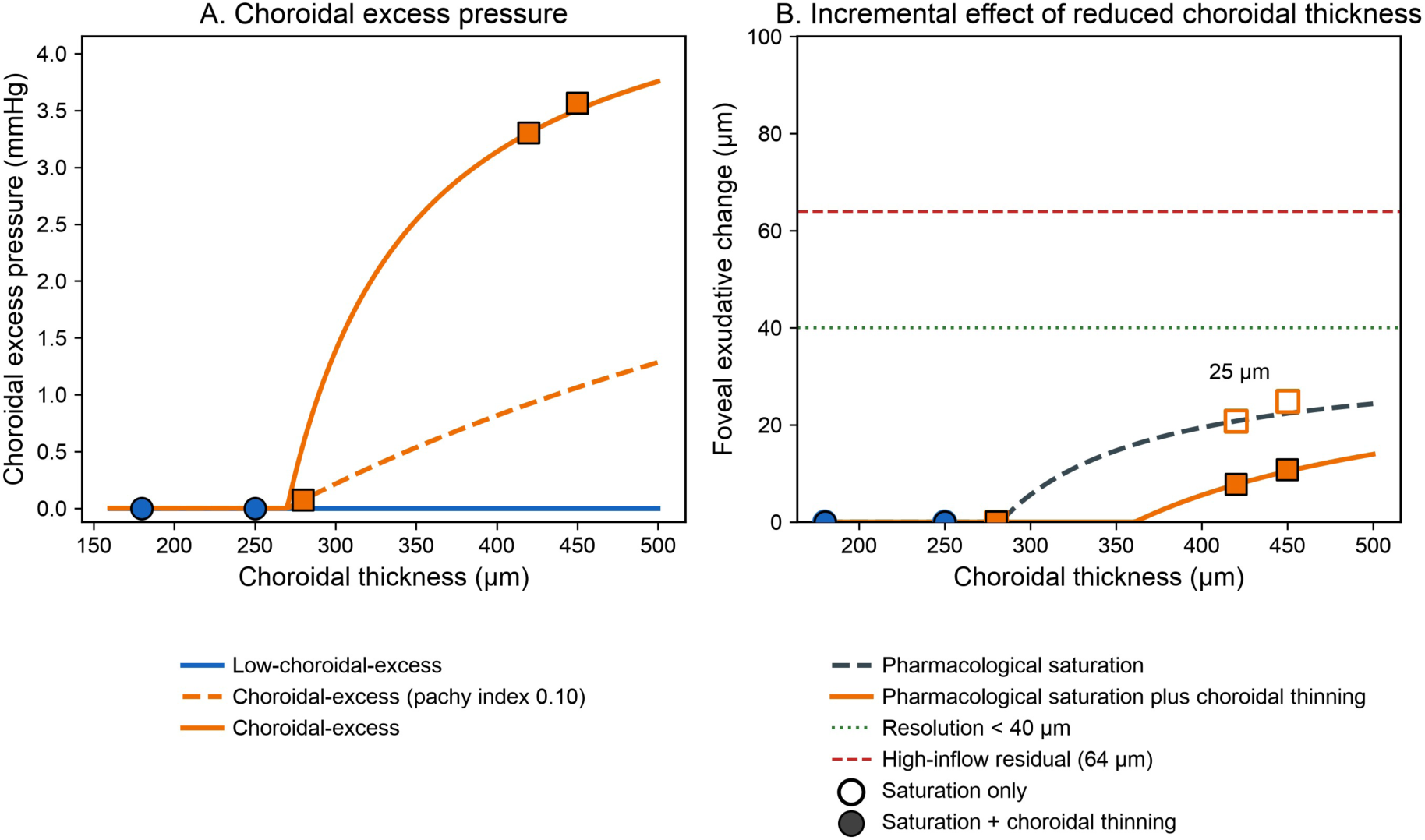
Choroidal excess pressure and the incremental effect of reduced choroidal thickness. A: Choroidal excess pressure versus choroidal thickness at pachy index 0, 0.10, and 0.85. B: Foveal exudative change after pharmacological saturation (dashed) and after the additional CT and pachy-index step of 0.81 and 0.55 from the published ratios [26] (solid). The open point at CT of 450 μm is about 25 μm after saturation. The green dotted line is 40 μm. The red dashed line is the 64 μm high-inflow residual under saturation, shown so that the 13–14 μm increment at CT of 450 μm can be compared with that larger residual.

The same CT reduction was subsequently applied under a condition in which choroidal excess pressure was already present. At a CT of 450 μm, approximately 25 μm of residual fluid remained after the permeability reduction described in Table 1. The anti-VEGF–associated reductions in CT and pachy-index then produced a further decrease of approximately 13–14 μm (Figure 3B). Thus, residual fluid changed with CT only in the presence of pre-existing choroidal excess pressure. Even under this condition, the magnitude of the reduction remained smaller than the residual fluid level observed under high inflow in the same panel (64 vs 13–14 μm).

### 3. Relative Rupture-Hazard Index

Reducing lesion pressure from high inflow to usual lesion pressure, with wall stability held constant, decreased the index to approximately 0.64 of the high-hazard reference. Wall stabilization was then added to that pressure reduction. Combining that pressure reduction with an increase in wall stability decreased the index to approximately 0.34 (Figure 4A). The corresponding exudative-change curves against effective transmural pressure are shown in Figure 4B. Open and filled points are the untreated and saturation outputs in Table 1. Black diamonds mark the Results 1 isolation from the choroidal-excess lesion pressure to high inflow (approximately 150 to 760 μm); the same pair at high-hazard permeability is approximately 17 to 311 μm. Rupture propensity fell when lesion pressure was lowered and fell further when the wall was also stabilized.

**Figure 4.**
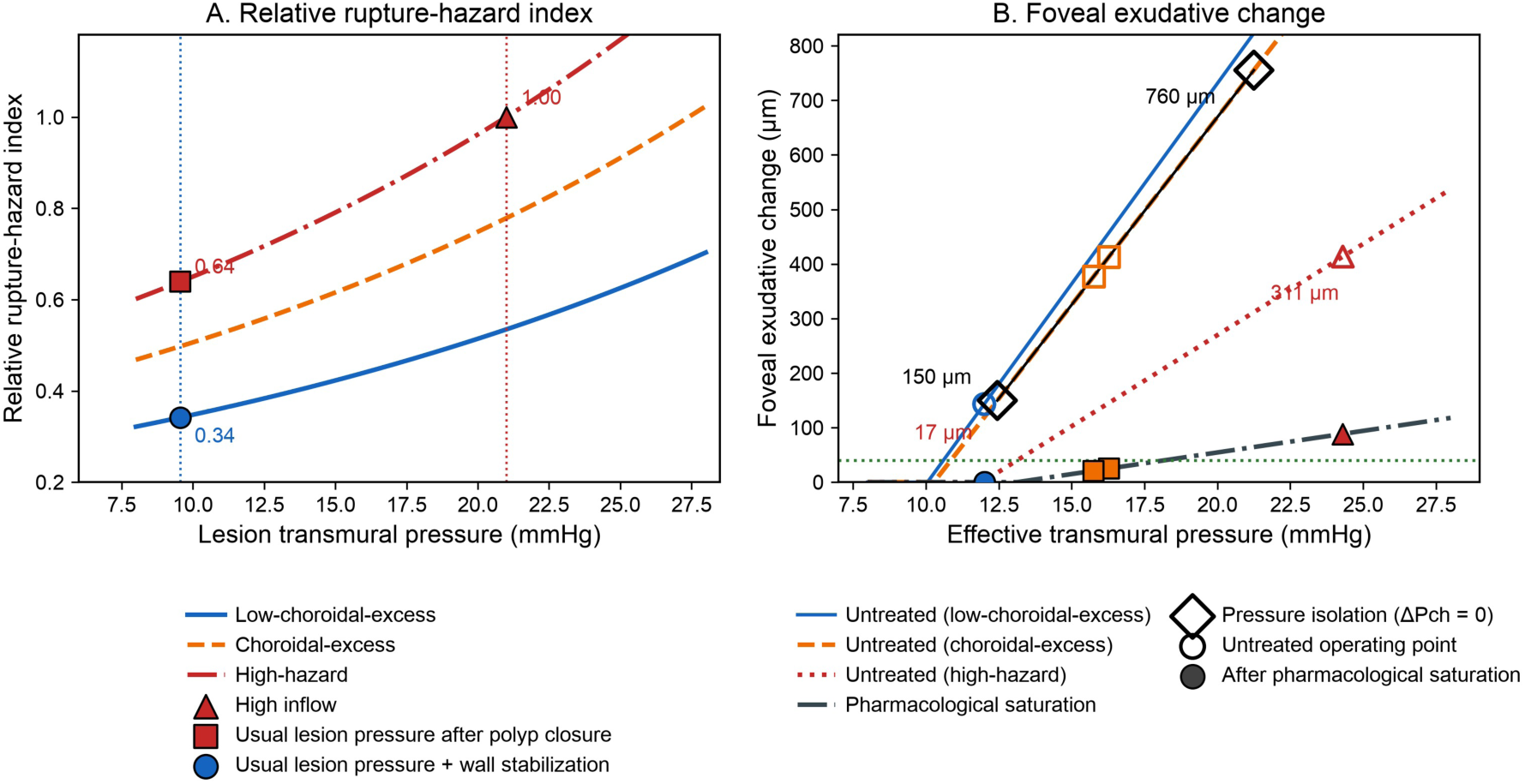
Relative rupture-hazard index against ΔPlesion and foveal exudative change against ΔPeff. A: Relative rupture-hazard index versus ΔPlesion at the wall-stability values of the low-choroidal-excess, choroidal-excess, and high-hazard states, scaled to 1.00 at high inflow. The vertical dotted lines are high inflow and usual lesion pressure. Three points mark the pressure-reduction sequence: high inflow (1.00), usual lesion pressure after a pressure reduction corresponding to polyp closure (0.64), and the same pressure with wall stabilization added (0.34). B: Foveal exudative change versus ΔPeff. Untreated curves are the low-choroidal-excess, choroidal-excess, and high-hazard permeabilities; the remaining curve is pharmacological saturation. Open and filled points are the untreated and saturation outputs in Table 1. Black diamonds, joined by a line, mark the Results 1 isolation at choroidal-excess permeability with choroidal excess set to zero (about 150 μm at the choroidal-excess lesion pressure and about 760 μm at high inflow). The same pair at high-hazard permeability is about 17 to 311 μm. The green dashed line is 40 μm.

### 4. Sensitivity Analyses

Table 2 summarizes the sensitivity analyses presented in Supplementary Figures 1 and 2. One-at- a-time analyses were used to rank the parameters that were varied. Residual thickness was primarily influenced by post-saturation permeability. The index following pressure reduction was influenced by the wall-thickness ratio and, when stabilization was included, by wall stability. At high inflow, the residual exudative change was evaluated across the Monte Carlo draws and remained above the resolution threshold throughout the sampled interval. The qualitative finding was robust, although the exact residual thickness was uncertain by approximately fourfold. This uncertainty arose because the reference condition assumes zero choroidal excess and therefore represents a lower-end estimate, with values sampled only upward from the permeability floor (Pbase=0.05).

**Table 2.** Sensitivity analysis (one-at-a-time ranges and Monte Carlo 95% intervals)

| Endpoint | Results / reference point | One-at-a-time range | Monte Carlo 95% interval (median) |
| --- | --- | --- | --- |
| 40-$\mu\text{m}$ boundary pressure (mmHg) | about 15 | about 10–18 | 9.1–17.3 (12.7) |
| Residual exudative change at high inflow, choroidal excess set to zero ($\mu\text{m}$) | 64 | 64–293 | 69–286 (170) |
| Relative rupture-hazard index after pressure reduction | about 0.64 | about 0.41–0.80 | about 0.41–0.78 (0.57) |
| Relative rupture-hazard index after pressure reduction plus wall stabilization | about 0.34 | about 0.22–0.64 | about 0.23–0.68 (0.39) |

The high-inflow working value and the tissue-pressure setting were then replaced in separate sensitivity analyses. Replacing the high-inflow working value with the diastolic value obtained using the human compression method or setting tissue pressure equal to the intraocular-pressure reference, resulted in a steady lesion pressure below the high-hazard cutoff (Supplementary Table S5 and Supplementary Figure 3). Finally, the resolution-threshold boundary was compared with the lesion pressure in the presence of choroidal excess. In 45% of the Monte Carlo draws, the simulated resolution-threshold boundary was at or below this pressure (Supplementary Figure 2A).

## Discussion

In the model, high inflow increased untreated exudative change when permeability was held constant, and residual exudative change after pharmacological saturation remained above the resolution threshold (Results 1; Figure 2). Sensitivity analyses supported the robustness of the qualitative conclusion that high inflow sustains a detectable residual exudative signal.

Incomplete responses to anti-VEGF therapy have been associated with arteriolarized or high-flow neovascular complexes [28][29]. Network vessels frequently fill concurrently with the surrounding choroidal arteries [12], whereas pulsatile polyps fill earlier and are associated with taller pigment epithelial detachments and a higher incidence of subsequent subretinal hemorrhage [11]. These findings are compatible with elevated lesion pressure. Hyaline degeneration of choroidal vessels, analogous to that seen in hypertensive arteries, has been well described [30][31]. However, pulsation is not specific to elevated lesion pressure. It may also occur in venous stasis, including caliber variation at arteriovenous crossings [32] and pulsatile filling of dilated choroidal veins across macular watershed zones [33]. Thus, pulsation alone cannot establish elevated lesion pressure from imaging findings. However, residual fluid after pharmacological saturation persisted in the elevated-lesion-pressure condition but not in the stagnation condition, suggesting that stagnation alone was insufficient within the modeled parameter space.

In the thickened choroidal-excess state, a reduction in CT and pachy-index of the magnitude reported after anti-VEGF treatment produced an additional decrease in exudative change after permeability reduction; this effect was absent when choroidal excess pressure was already near zero (Results 2; Figure 3). This additional decrease is consistent with choroidal venous congestion, which underlies the venous-overload hypothesis of increased transmural driving pressure [8]. Following anti-VEGF treatment, both subfoveal choroidal thickness and the choroidal vascularity index have been reported to decrease [26], whereas eyes with choroidal thickening may show a less favorable therapeutic response [34]. ΔPch represents the contribution of choroidal venous overload to the increase in driving pressure. However, even in the thickened choroidal-excess state, ΔPch remained only a few mmHg (Table 1). Thus, this choroidal component alone could not account for the larger increase in driving pressure. When compared directly, the larger pressure increment was attributable to high inflow rather than choroidal thickening itself.

Lowering lesion pressure from high inflow to usual lesion pressure decreased the index to approximately 0.64 of the high-hazard reference, and to approximately 0.34 when wall stability was also increased (Results 3; Figure 4). Fluid resolution, however, represents a distinct outcome. A dry OCT does not necessarily indicate a lower rupture index, and persistent polyps after fluid resolution may indicate persistence of either a pressure-related or wall-stability component.

Consistent with this interpretation, polyp regression has been associated with a lower subsequent risk of massive submacular hemorrhage (SMH) [25]. This distinction is also reflected in clinical trial data, in which resolution of exudative activity was more frequent than complete polyp regression. In PLANET, complete polyp regression at week 52 was observed in 38.9% of eyes, whereas 81.7% had no polypoidal lesions with leakage [22]. Similarly, in SALWEEN, complete polypoidal lesion regression at week 48 occurred in 60.8% of eyes, whereas 86.0% had inactive polyps [35]. Thus, exudative activity may resolve without complete polyp regression, a dissociation that is compatible with, but does not establish, persistence of pressure-related or wall-stability components after fluid resolution.

## Limitations

This study has several limitations. First, prospective patient-level validation remains to be done. Second, the four states are ordered regions of a continuous space rather than a clinical decision rule. Lesion pressure and wall stability were mapped jointly from a single PCV-versus-typical-nAMD comparison [17], so those components are not separated. Wall fatigue remains outside the model. The published post–polyp-regression hazard ratio of 0.271 came from the same institution as that comparison, and cohort overlap remains possible [25].

ΔPch, the additive form of ΔPeff, and the CT-reduction step are model constructions rather than measurements: ΔPch is generated from CT and the pachy index, ΔPeff is additive by assumption [7][8], and the CT scenario follows published anti-VEGF reductions in one study [26]. Where lesion transmural pressure is Windkessel-derived, outflow resistance contributes both to lesion transmural pressure and, through the multiplier f, to ΔPch, so this form may overrepresent resistance-related effects. Choroidal autoregulation and local vascular architecture are more complex than the lumped-parameter representation [36–39]. Sensitivity analysis demonstrated that the absolute magnitude of residual thickness and the position of the resolution-threshold boundary remained sensitive to uncertainty in post-saturation permeability and reference-state assumptions.

What remains for a patient-level test is a clinical course and a measurement set. PCV can pass from an exudative, thick-choroid phase to polyp pulsation, enlargement of pigment epithelial detachment, or SMH at a later time [11][27]. Morphologic analysis of MNV on OCTA [40], with retinal fluid, choroidal thickness, and choroidal vascularity index, could separate lesion-side mechanical stress from wall stability. Later massive SMH and treatment burden are the events against which those quantities can be scored.

## Conclusions

Exudation and rupture propensity were not interchangeable outputs. In the model, saturation reduced fluid. Residual fluid after saturation, and an incremental decrease when choroidal thickness was reduced in thickened states, support two imaging hypotheses—pressure-related residual fluid, and possible discordance between fluid resolution and rupture-related risk—that require patient-level testing. Lowering lesion pressure lowered the rupture index.

## Supporting information

Video 1. Effective transmural pressure over the cardiac cycle

## Data Availability

The computational code supporting the findings of this study is available to reviewers and from the corresponding author on request until a public version-tagged archive is deposited.

## Ethics

This study was a theoretical mathematical analysis and did not involve human participants, identifiable patient information, or a clinical intervention. Institutional review board approval and informed consent were not applicable.

## Data and code availability

No individual-participant data were used in this study. The computational code supporting the findings of this study is available to reviewers and from the corresponding author on request until a public version-tagged archive is deposited.

## Acknowledgments

A generative artificial intelligence tool (Cursor, version 3.20.17; Anysphere, Inc., San Francisco, CA, USA) was used to assist with English language editing, manuscript formatting, checking of citations, and generation of figures. The authors reviewed and verified all text, numbers, equations, references, and figures, and take full responsibility for the content of the manuscript.

---

**Video 1. Variation of effective transmural pressure over the cardiac cycle in the four states. Each panel is a state in Table 1. The solid line is ΔPeff; the dashed line is ΔPlesion alone. The residual-exudation and high-hazard states are pressure intervals, not single points. Oscillation is the Windkessel response to arterial pulsation of 22 mmHg (60 beats/min).**

## Supplementary Material

Supplementary Tables S1–S8 and Supplementary Figures 1–5. Main-text reference numbers [1]–[40] are retained. Sources cited only here are listed at the end as [S1]–[S18].

This file contains the shared model inputs; the permeability signaling equations; the exploratory drug-parameter analysis; extended sensitivity methods and results; alternative rupture-hazard functional forms; the 0.271 recalibration; and replacement of the 51-mmHg reference for high inflow.

**Supplementary Table S1.**
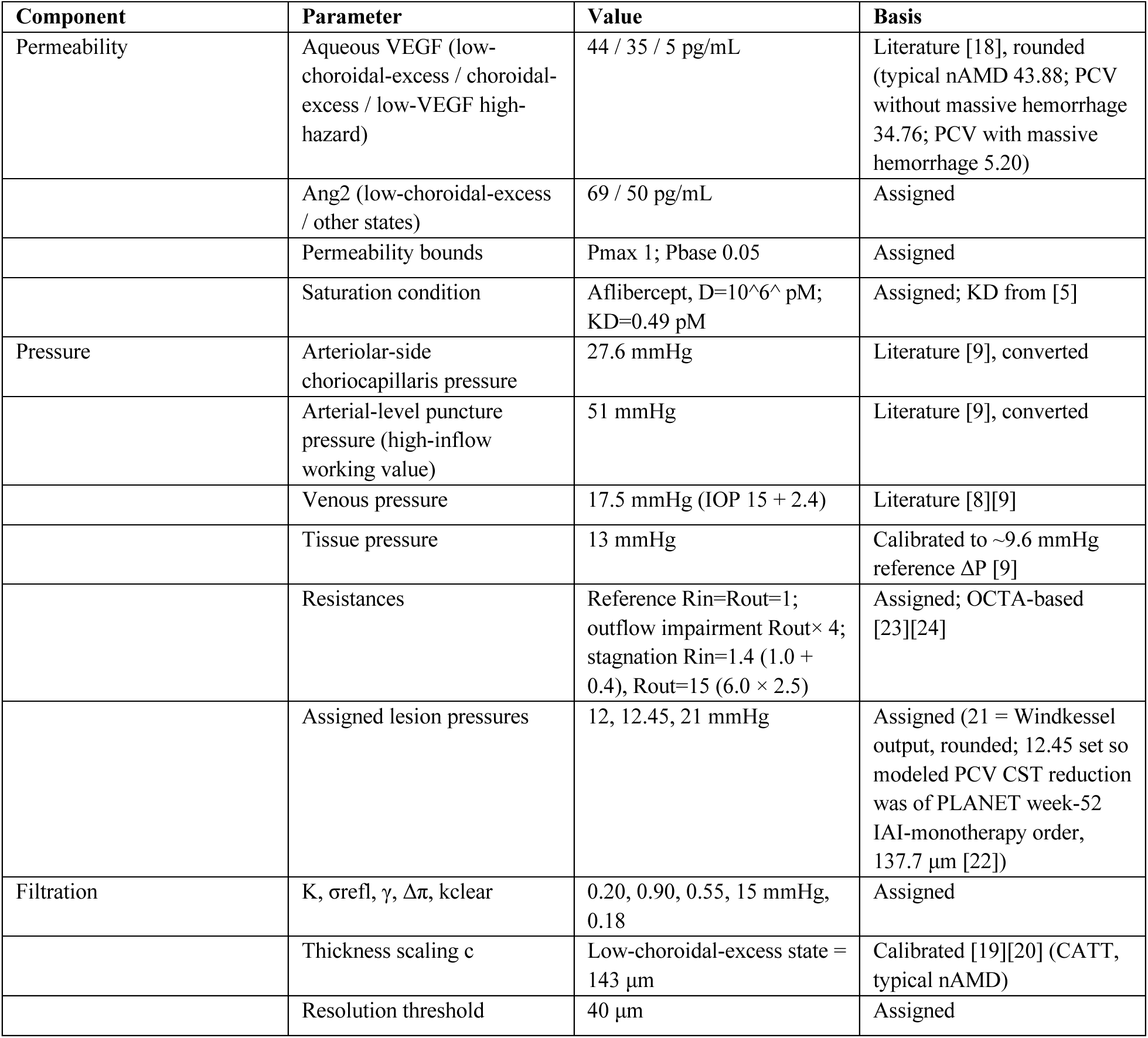
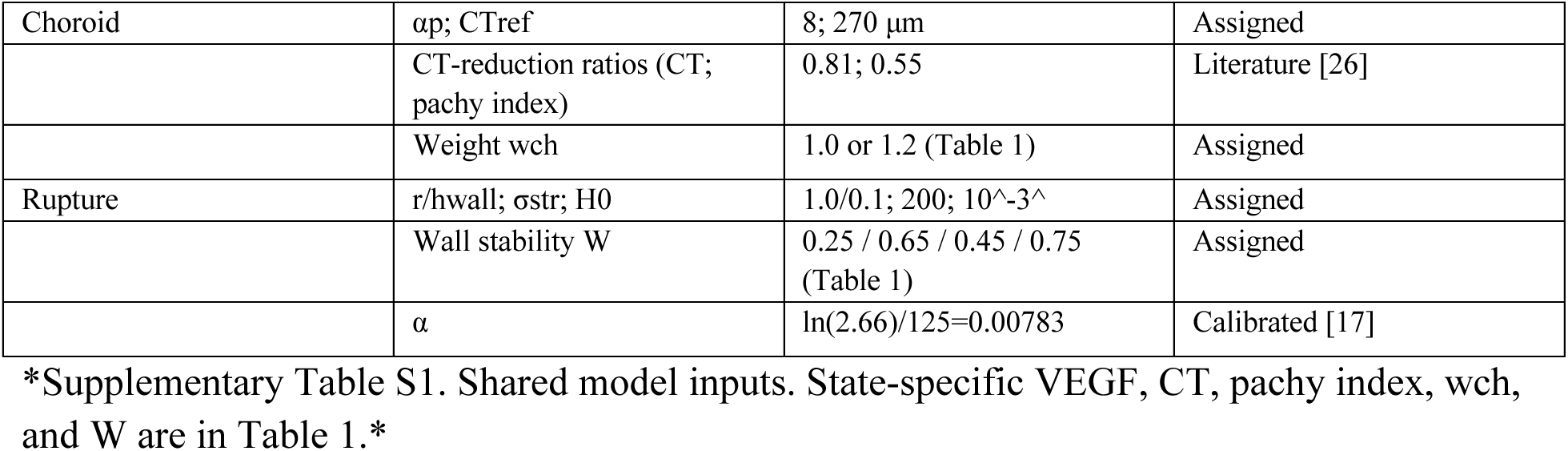
Model inputs. “Literature” = taken from the cited source; “Assigned” = chosen for the model; “Calibrated” = set to match a cited quantity. This table is the numerical-input list for the main-text Methods. State-specific values are in Table 1; signaling constants are in Supplementary Table S2.

**Supplementary Table S2.** Constants of the permeability model.

kV, kT, $\delta$ S, kP, and $\delta$ P are coefficients assigned as relative magnitudes of Src activity and vascular permeability.
| Symbol | Value | Unit / meaning |
| --- | --- | --- |
| kV | 1.0 | Src activation by VEGF |
| KV,1/2 | 50 | VEGF half-saturation (same units as aqueous concentration) |
| kT | 1.5 | Src inactivation by Tie2 |
| δS | 0.3 | Basal inactivation of Src |
| kP | 1.0 | Permeability from Src |
| δP | 0.5 | Relaxation of permeability |
| Pmax | 1.0 | Upper bound of permeability |
| Pbase | 0.05 | Lower bound of permeability |
| A1 | 100 | Ang1 (relative amount) |
| A2 | 69 (low-choroidal-excess state) / 50 (other states) | Ang2 (same units as aqueous concentration) |
| KT,1/2 | 100 | Ang1 half-saturation of Tie2 |
| KI | 100 | Ang2 inhibition constant |
| KD,Ang2 | 100 nM (main analysis) / 22 nM (faricimab) | Anti-Ang2 dissociation constant [6] |

When an anti-Ang2 agent was included, effective Ang2 was $A2/(1+D_{Ang2}/KD_{Ang2})$ . Tie2 activity T, Src activity S, and permeability P were

**Supplementary Table S3.**
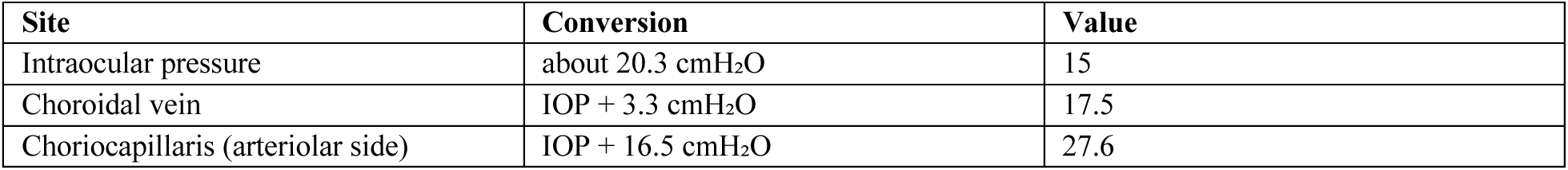
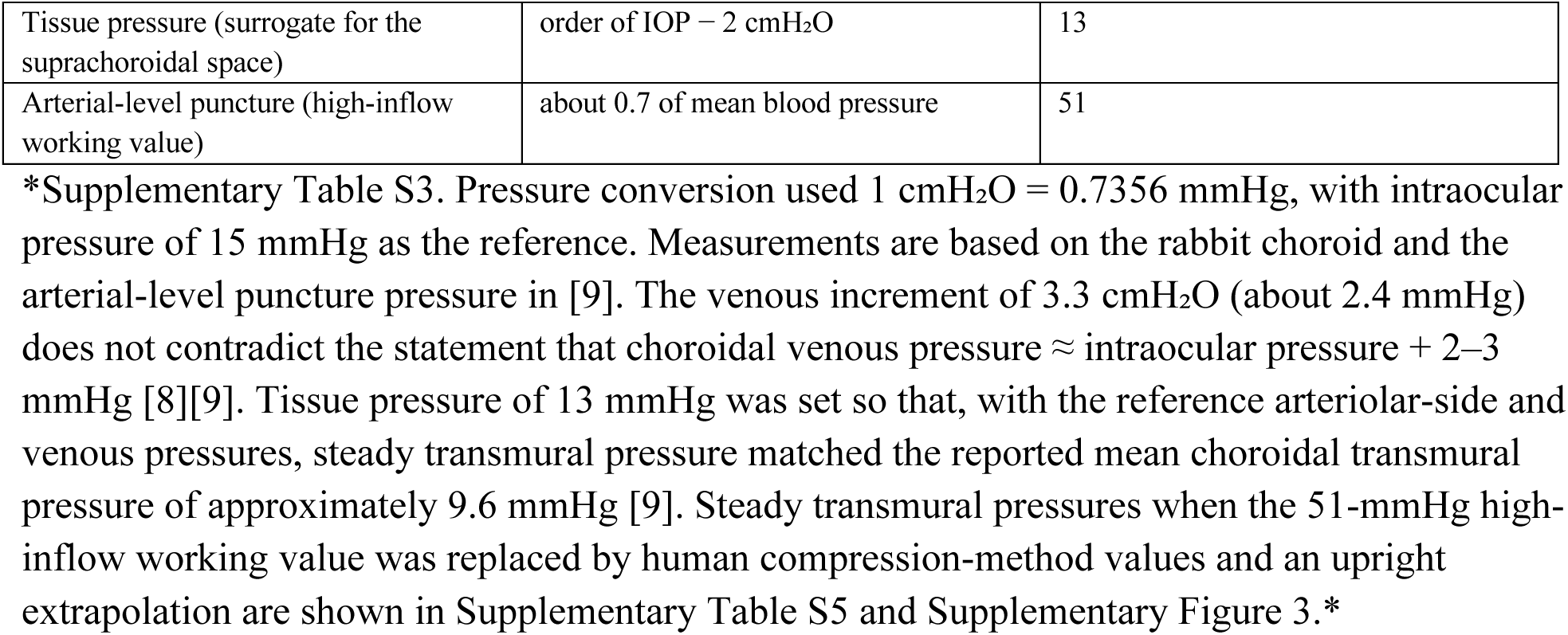
Pressures used in the conversion (mmHg) [9].

**Supplementary Table S4.** Human compression-method arterial pressures.

| Subjects | Site | Systolic (mmHg) | Diastolic (mmHg) |
| --- | --- | --- | --- |
| Normal (9 cases) | Nasal | 62 (59, 67) | 41 (39, 47) |
| Normal (9 cases) | Temporal | 57 (56, 59) | 37 (31, 43) |
| All eyes (n = 40) | Nasal | 51 (45, 63; 39 eyes) | — |
| All eyes (n = 40) | Temporal | 51 (44, 57) | — |

\*Supplementary Table S4. All values were obtained by compressing a vessel on the ocular surface and taking appearance of the pulse or collapse of the vessel as the endpoint; they are a measurement separate from puncture pressures in the monkey [9][36][37].
| Subjects | Systolic (mmHg) | Diastolic (mmHg) | Arterial diameter (μm) |
| --- | --- | --- | --- |
| Normal, right (9 eyes) | 59.2 (58.2, 63.5) | 35.2 (28.6, 38.3) | 61.2 (58.5, 71.2) |
| Normal, left (8 eyes) | 59.4 (56.0, 60.3) | 31.5 (28.5, 35.7) | 55.3 (50.4, 64.0) |
| Ocular hypertension, right (29 eyes) | 55.6 (50.2, 62.8) | 30.6 (26.0, 35.3) | 77.2 (72.8, 82.0) |
| Ocular hypertension, left (29 eyes) | 55.3 (46.7, 60.5) | 28.3 (23.8, 35.8) | 71.3 (65.3, 81.3) |

**Supplementary Table S5.**
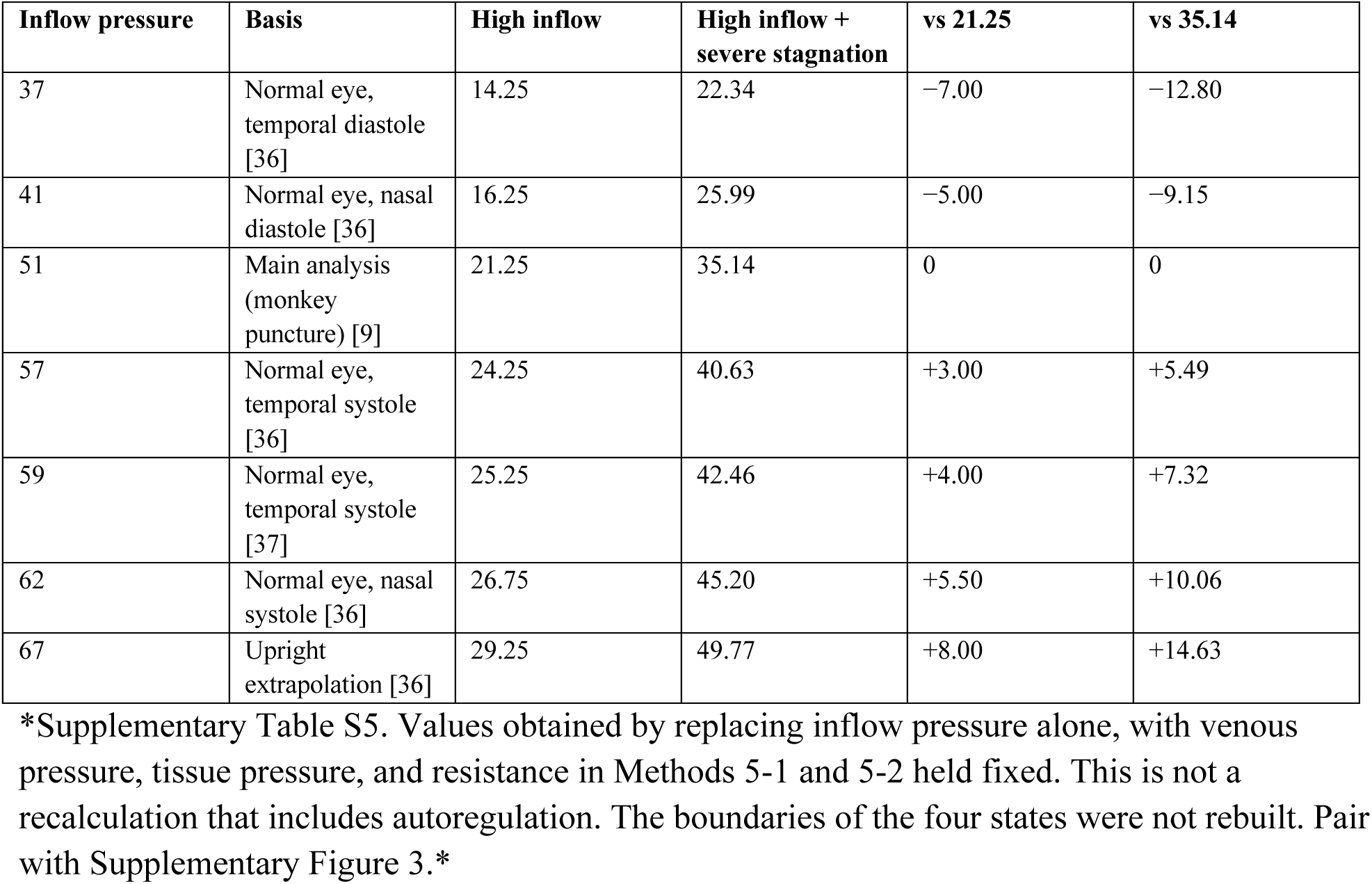
Steady transmural pressure (mmHg) after replacement of the inflow pressure in Supplementary Table S3.

**Supplementary Table S6.** Comparison of functional forms and validity against a separate cohort from the same institution.

The main analysis is the exponential form.
| Functional form | Pressure reduction alone | Pressure reduction + wall stabilization |
| --- | --- | --- |
| Exponential (main analysis) | about 0.64 | about 0.34 |
| Linear (stress difference) | about 0.71 | about 0.31 |
| Power of the stress ratio | about 0.54 | about 0.31 |

### Exploratory drug-parameter scenario analysis

This analysis examined modeled consequences of selected drug-related parameters after three loading injections on days 0, 28, and 56, with simulation through day 360. It was not designed to compare clinical efficacy, durability, or treatment intervals.

Intravitreal concentration was modeled as first-order decay D(t)=D0 2^(-t/t1/2)^. At each time point the concentration was substituted into the permeability equation. Threshold-crossing day was the first day after three injections on which modeled foveal exudative change again reached or exceeded 40 μm. Separately, the single-dose time at which free VEGF exceeded 1% of total VEGF was

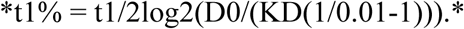

Assigned immediate post-injection concentrations were 10^6^ pM for ranibizumab, aflibercept 2 mg, and faricimab; 4×10^6^ pM for aflibercept 8 mg; and 2.215×10^7^ pM for brolucizumab, from the molar-dose ratio (6/26)/(0.5/48)=22.15 [S1–S5]. Biosimilars used the reference-molecule constants [S1][S2]. CT and pachy-index ratios (Supplementary Table S7) followed the reported rank order of SFCT and CVI reductions for ranibizumab, aflibercept, and faricimab [26]; the aflibercept 8-mg scenario used the 2-mg ratios; brolucizumab used a 1-month CT ratio of 196.0/231.9 μm (0.85) and a pachy-index ratio of 0.80 [S6]. CT and the pachy index took their post-remodeling values from day 56 onward. In the low-choroidal-excess state, ΔPch was 0.

Sensitivity repeats used an aqueous half-life of 9 days for brolucizumab [S7] and 15 days for aflibercept 8 mg [S8].

### Results

Single-dose times to 1% free VEGF were 61 days (ranibizumab), 130 days (aflibercept 2 mg), 149 days (aflibercept 8 mg), 76 days (faricimab), and 62 days (brolucizumab). After three injections, threshold-crossing days in the low-choroidal-excess state were 166, 241, 260, 191, and 150; in the choroidal-excess state they were 128, 198, 216, 145, and 127. Incremental decreases in the choroidal-excess CT scenario were approximately 3, 6, 6, 14, and 8 μm (Supplementary Table S8 and Supplementary Figure 5). The ranibizumab biosimilar scenario matched the reference ranibizumab scenario. Under combined high inflow and choroidal thickening, residual modeled exudative change was approximately 90 μm across drug-parameter scenarios.

A 9-day aqueous half-life for brolucizumab shifted the low-choroidal-excess threshold-crossing day to 227 days. A 15-day half-life for aflibercept 8 mg kept modeled suppression beyond day 360.

These outputs follow the assigned concentrations, half-lives, and CT ratios. They are not clinical recurrence times, dosing intervals, or a ranking of agents [S3][S5][S9–S11].

**Supplementary Table S7.** Drug-related input parameters used in exploratory model scenarios.

Ratios for ranibizumab, aflibercept, and faricimab follow the reported SFCT/CVI order [26]. The brolucizumab CT ratio is from a 1-month post-switch report [S6].\*
| Drug | VEGF KD (pM) | Intravitreal half-life (days) | Immediate post-injection concentration ratio | Choroidal-thickness ratio | Pachy-index ratio |
| --- | --- | --- | --- | --- | --- |
| Ranibizumab | 46 | 7.9 | 1 | 0.93 | 0.92 |
| Ranibizumab | 46 | 7.9 | 1 | 0.93 | 0.92 |
| biosimilar |  |  |  |  |  |
| Aflibercept 2 mg | 0.49 | 9.1 | 1 | 0.88 | 0.80 |
| Aflibercept 8 mg | 0.49 | 9.1 | 4 | 0.88 | 0.80 |
| Faricimab | 46 (VEGF),<br>2.2×10<sup>4</sup> (Ang2) | 9.8 | 1 | 0.81 | 0.55 |
| Brolucizumab | 28.4 | 4.8 | 22.15 | 0.85 | 0.80 |

**Supplementary Table S8.** Exploratory outputs after a fixed three-injection loading schedule.

Threshold-crossing day is the first day after injections on days 0, 28, and 56 on which modeled foveal exudative change again becomes $\geq 40 \mu\text{m}$ .
| Drug | Modeled days to 1% free VEGF (single dose) | Threshold-crossing day, low-choroidal-excess | Threshold-crossing day, choroidal-excess | Incremental CT-scenario decrease, choroidal-excess |
| --- | --- | --- | --- | --- |
| Ranibizumab / biosimilar | 61 | 166 | 128 | about 3 $\mu\text{m}$ |
| Aflibercept 2 mg | 130 | 241 | 198 | about 6 $\mu\text{m}$ |
| Aflibercept 8 mg | 149 | 260 | 216 | about 6 $\mu\text{m}$ |
| Faricimab | 76 | 191 | 145 | about 14 $\mu\text{m}$ |
| Brolucizumab | 62 | 150 | 127 | about 8 $\mu\text{m}$ |

**Supplementary Figure 5.**
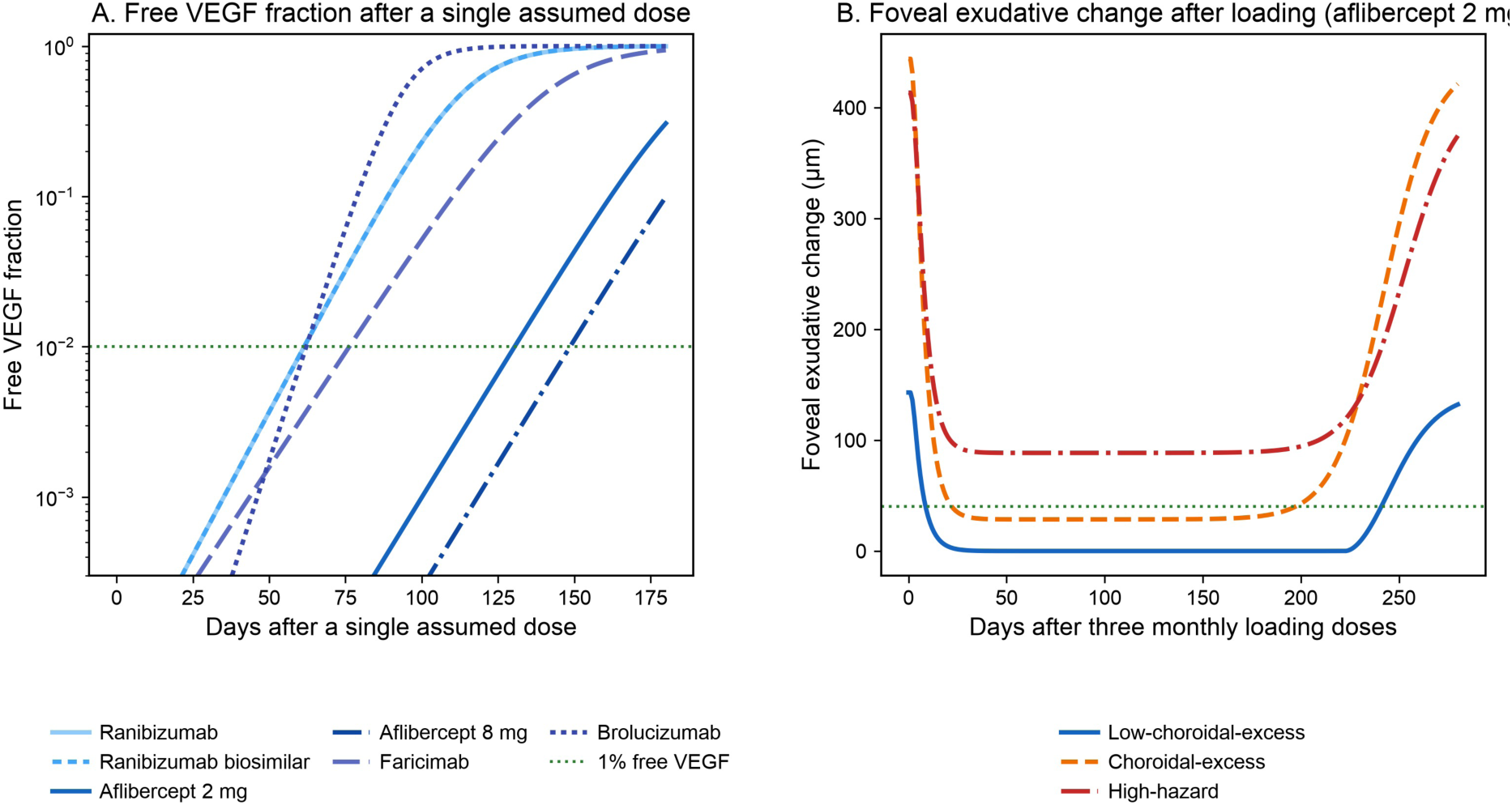
Time course of the free-VEGF ratio and of modeled foveal exudative change after assumed anti-VEGF administration. A: Free-VEGF ratio after an assumed single dose. The green dashed line is 1%. B: Modeled foveal exudative change after three monthly injections of aflibercept 2 mg. Blue, orange, and red are the low-choroidal-excess, choroidal-excess, and high-hazard states. Under high inflow the value exceeds 40 μm throughout.

### Extended sensitivity methods

Parameters were varied individually across predefined ranges, and 4000 independent sets were drawn from uniform distributions over the same ranges. Post-saturation permeability was sampled from Pbase=0.05 to 0.20; the lower bound is the saturation floor in Supplementary Table S2. The other sampled ranges were lesion pressure 8–35 mmHg, choroidal thickness 250–500 μm, αp 4–12, wch 0.5–1.5, wall stability 0.15–0.75, wall ratio r/h 5–20, and the 40-μm criterion 30–60 μm. The analyses retained the four-state boundaries, the literature values used for calibration [17][19][20][22], the CT and pachy-index ratios in Supplementary Table S7, the drug-concentration time-course assumptions, and α.

Two alternative rupture-hazard forms were calibrated to the same two reference points as the exponential: a linear function of wall-stress difference, and a stress-ratio function (σwall/σmax)^β^ with β=0.78 [15][16]. The published hazard ratio of 0.271 after polyp regression [25] was used as an external descriptive comparator. Three conditions were compared: no regression (21 mmHg; wall stability, 0.25); pressure reduction at the reference pressure with wall stability 0.25; and the reference pressure with wall stability 0.75. In one analysis the exponential was recalibrated so that the combined condition equaled 0.271, after which the high-hazard-to-low-choroidal-excess ratio was calculated.

The 51-mmHg high-inflow working value in Supplementary Table S3 was replaced with human compression-method estimates (Supplementary Table S4), approximately 37–62 mmHg from diastole to systole in normal eyes, and with an upright extrapolated value of 67 mmHg [36][37]. Venous pressure, tissue pressure, vascular resistance, and the resistance set for severe stagnation were unchanged. Tissue pressure was not among the one-at-a-time or Monte Carlo parameters.

### Extended sensitivity results

Under pressure reduction alone, the linear and stress-ratio forms yielded relative indices of approximately 0.71 and 0.54; with wall stabilization both yielded approximately 0.31 (Supplementary Table S6 and Supplementary Figure 4). Recalibration of the exponential to 0.271 gave a high-hazard-to-low-choroidal-excess modeled ratio of approximately 3.3.

The faricimab CT scenario produced the largest incremental modeled decrease in 70% of Monte Carlo draws (Supplementary Figures 1 and 2). When ΔPlesion was varied over 8–35 mmHg, residual modeled exudative change ranged from 0 to 583 μm and the relative index after pressure reduction ranged from approximately 0.20 to 1.0.

Replacing the 51-mmHg reference for high inflow with 37–67 mmHg yielded the steady lesion pressures in Supplementary Table S5 and Supplementary Figure 3. At 37 mmHg (diastolic value in the cited human measurements), lesion transmural pressure at high inflow was 14.25 mmHg and lay below the 21-mmHg cutoff. Systolic values of 57–62 mmHg and the 67-mmHg upright extrapolation exceeded 21 mmHg. When tissue pressure was set equal to the 15-mmHg intraocular-pressure reference, with venous pressure and inflow unchanged, lesion transmural pressure at high inflow was 19.25 mmHg, below the 21-mmHg cutoff. The four-state boundaries were not re-estimated. The value in the choroidal-excess row of 12.45 mmHg was at or above the simulated 40-μm boundary in 45% of Monte Carlo draws (median boundary, 12.7 mmHg).

### Supplement. Indirect descriptions of pressure in polyps

None of the following is evidence that polyps carry pressure equivalent to the high-inflow working value.

-Pulsatile filling of choroidal veins that cross a watershed is described as a venous-side pressure gradient [S12]. - Descriptions of dilation of the branching vascular network and its margin contain no numerical lesion pressure [S13]. - Choroidal neovascularization is less frequent when a cilioretinal artery is present [S14]. - Lower choroidal blood flow in the fellow eye is associated with progression to neovascularization [S15].

### Supplement. Autoregulation of choroidal blood flow

Subsequent work organizes choroidal blood flow by the difference between mean arterial pressure and intraocular pressure, rather than a fixed arterial-level working value, and states that flow is autoregulated down to a moderate fall in perfusion pressure [38][39]. This paper uses fixed boundaries of the representative condition; time variation of mean arterial pressure and intraocular pressure, and the threshold band of autoregulation, were not calculated.

Supplementary Table S5. is steady transmural pressure after replacement of inflow pressure alone, not a recalculation that includes autoregulation.

### Supplement. Detail moved from Limitations

The slope of the peak wall rupture index of abdominal aortic aneurysm was not used as the coefficient of rupture hazard, because the units and geometry differ. The aneurysm literature uses a stress-to-strength ratio; the exponential of a stress difference in the main analysis is a modeling choice [15][16]. Mapping post-regression to the reference pressure and the vessel wall of the low-choroidal-excess state is an assumption.

A difference in the sign of CVI shown in another report was not used for the choroidal-thickness ratios [S16]. The report that faricimab was least frequent as the last injection is a composition ratio, not an absolute incidence [S17]. The VEGF-side KD of faricimab depends on the assay system; under the saturation approximation it is of the same order as ranibizumab. The KD of brolucizumab is a KinExA value and is on a different measurement scale from surface plasmon resonance used for the other agents. The initial concentration of brolucizumab is a relative value obtained by multiplying the reference concentration assigned to ranibizumab by the molar ratio, not a conversion to absolute concentration that uses vitreous volume.

The time course of structural leakage in MNV, intraocular inflammation, the incidence of SMH, and fitting of a Weibull distribution to 10-year cumulative incidence were not examined. The low-choroidal-excess state is a state in which choroidal excess pressure is nearly 0 and exudation is determined mainly by vascular permeability; it does not mean that advanced type 3 MNV is discontinuous with the choroidal circulation [1][2][S18].

## Supplementary figure legends

**Supplementary Figure 1.**
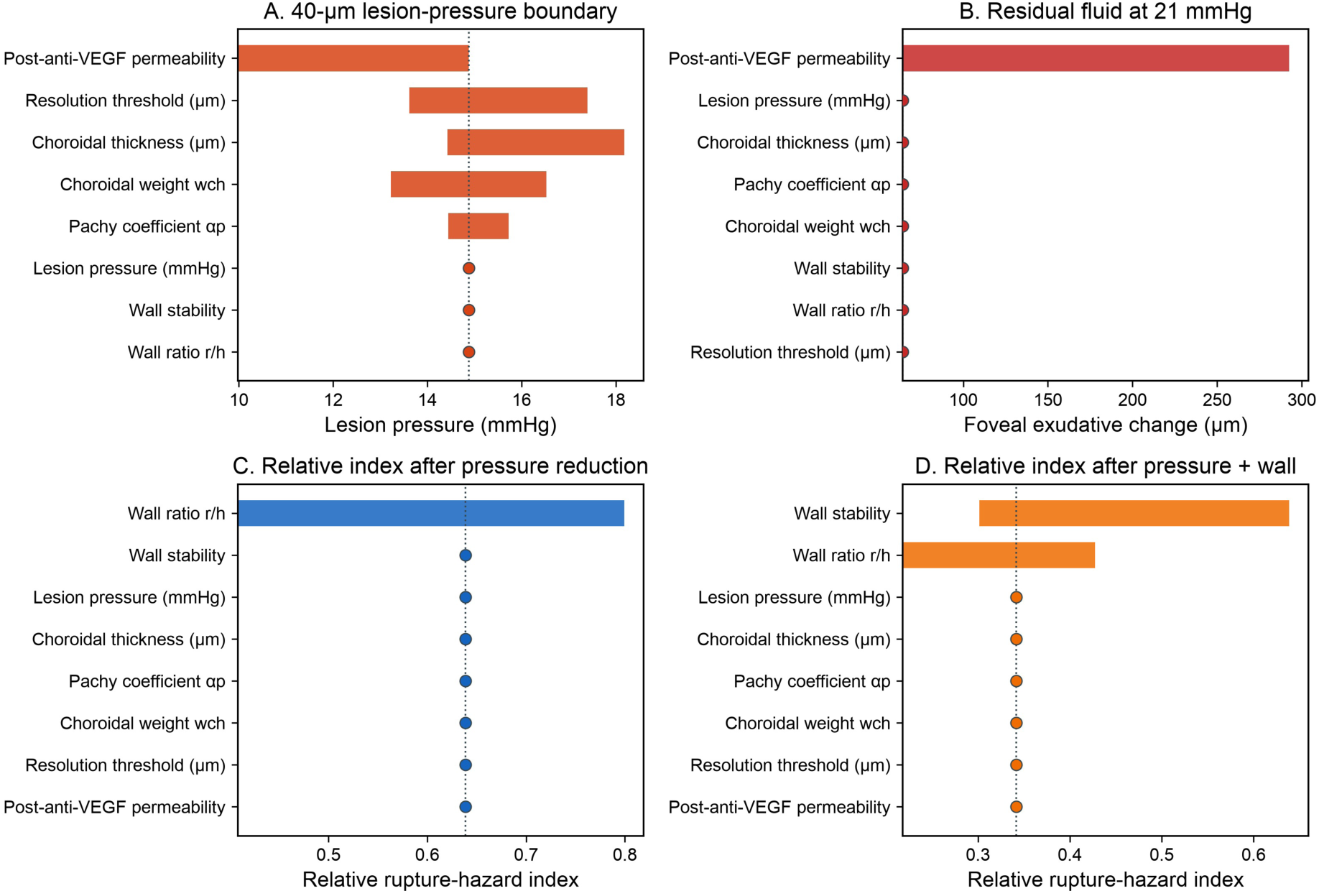
One-at-a-time sensitivity analysis. Each bar is the minimum to maximum when that parameter alone is moved across its predefined range. The dotted line is the reference-case value. A: 40-μm boundary pressure. B: Residual modeled exudative change under high inflow (21 mmHg) with choroidal excess pressure of 0. C: Relative rupture-hazard index after pressure reduction (reference about 0.64). D: The same index with wall stabilization (reference about 0.34). Values are the reduced index divided by the high-hazard reference, as in Table 2.

**Supplementary Figure 2.**
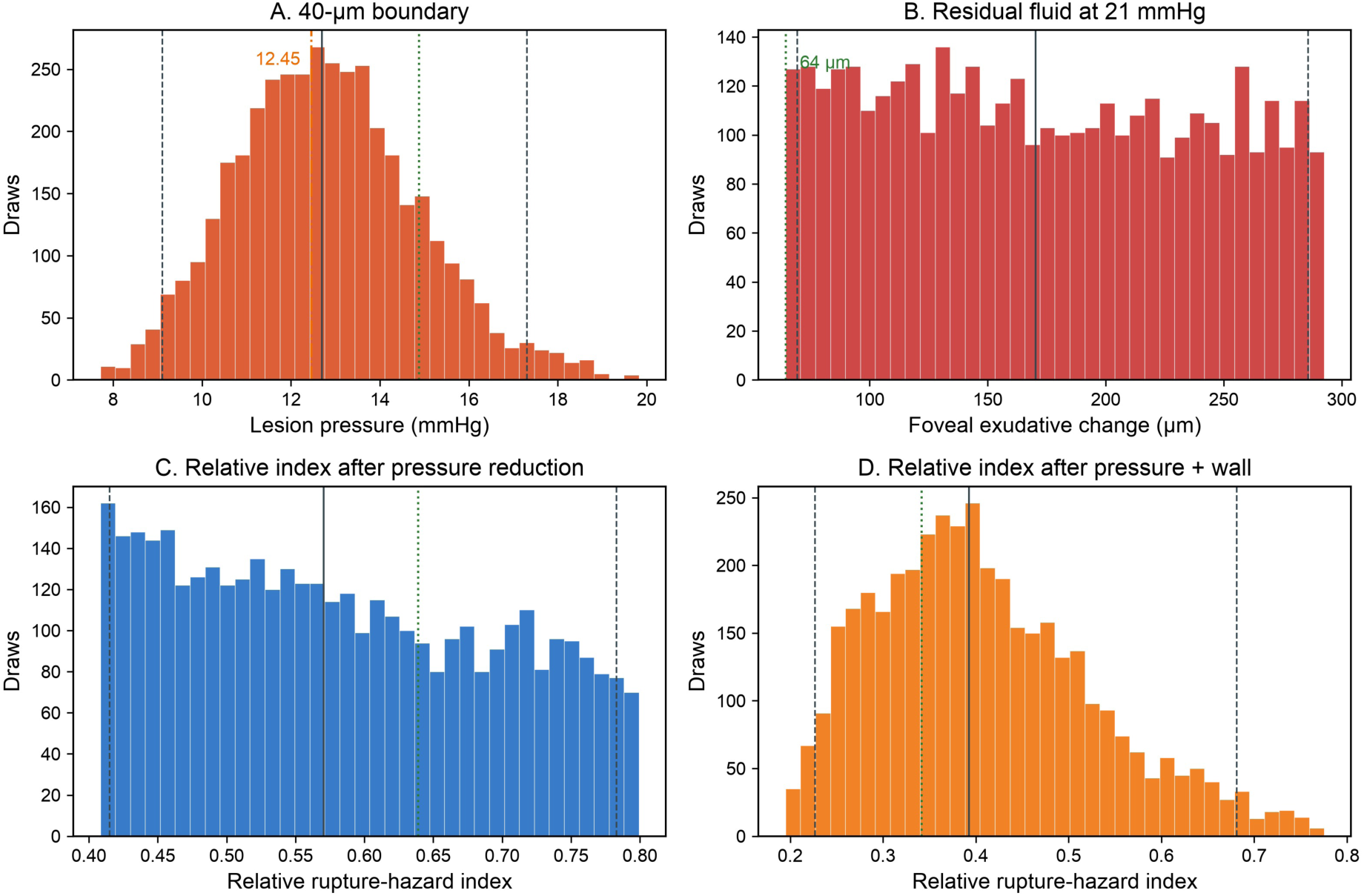
Monte Carlo 95% intervals. Four thousand draws from uniform distributions over the same ranges. Solid line, median; dashed lines, 2.5th and 97.5th percentiles; green dotted line, reference-case value. The orange dash-dotted line in A is the choroidal-excess lesion pressure. In B the green dotted line is the 64 μm lower-end reference, to the left of the Monte Carlo interval. Panels as in Supplementary Figure 1. Panels C and D use the same relative-index orientation as Table 2 (about 0.64 and 0.34).

**Supplementary Figure 3.**
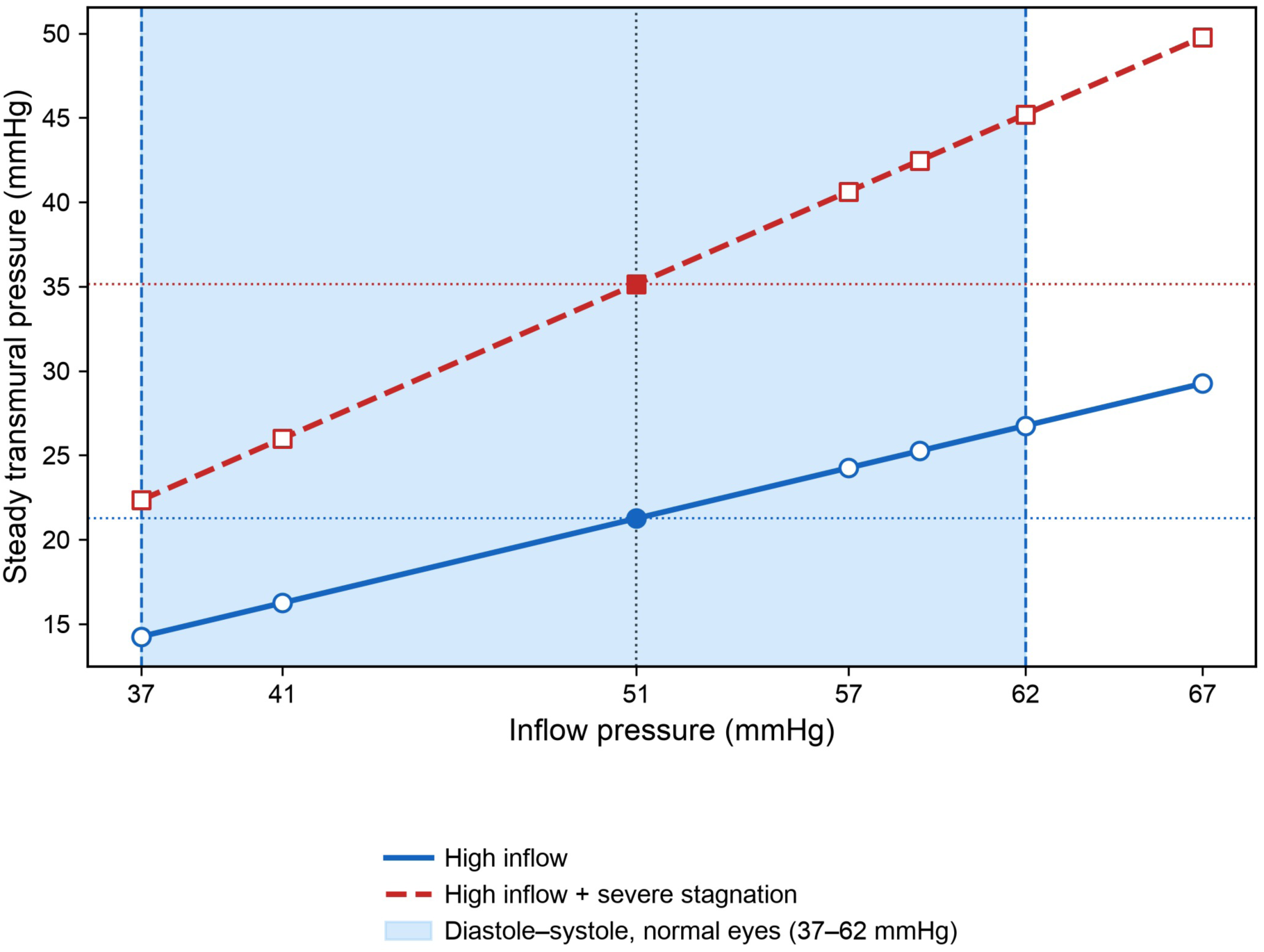
Steady transmural pressure against the inflow pressure in Supplementary Table S3. The solid line is high inflow; the dashed line adds severe stagnation. The vertical dotted line is the 51-mmHg main-analysis reference. The colored band is diastole to systole in normal eyes (about 37–62 mmHg). The 67 mmHg point is an upright extrapolation [36][37].

**Supplementary Figure 4.**
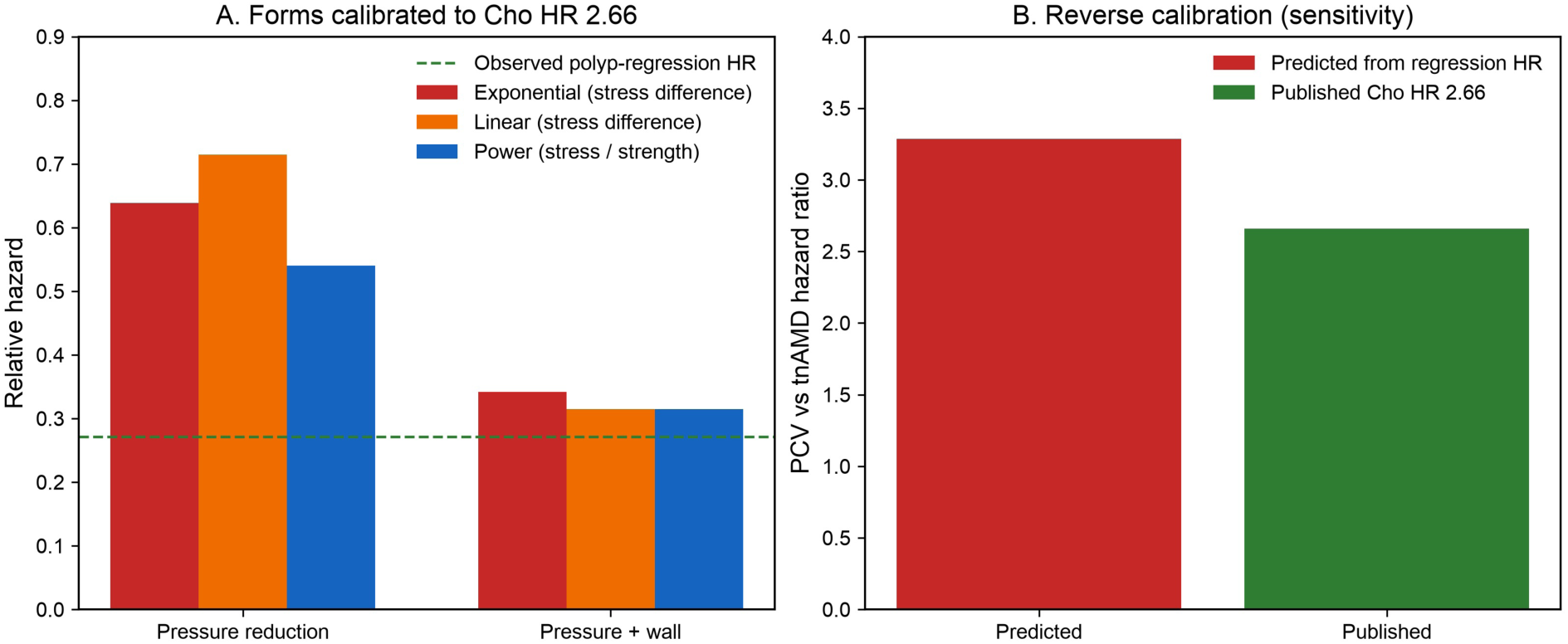
Functional form of the relative rupture-hazard index and comparison with a separate cohort from the same institution. A: Three functional forms fixed at the two points of the published hazard ratio 2.66. Left, pressure reduction alone; right, pressure reduction plus wall stabilization. The green dashed line is the post–polyp-regression hazard ratio of 0.271 [25]. B: Modeled high-hazard-to-low-choroidal-excess ratio (left) and the published value 2.66 (right) after recalibration of the exponential to 0.271.

