## Supplementary figures and images for "Four-State Hemodynamic Framework for Neovascular Age-Related Macular Degeneration: Mathematical Model Integrating Vascular Permeability, Lesion Transmural Pressure, Choroidal Excess Pressure, and Wall Stability"

### Video 1. Effective transmural pressure over the cardiac cycle

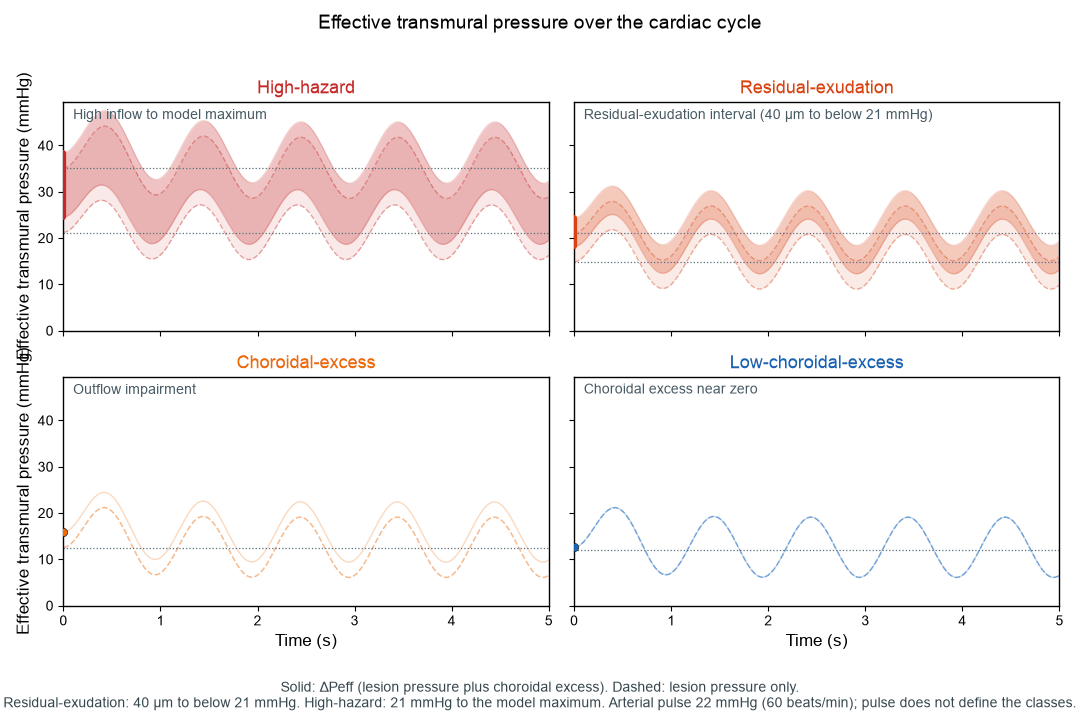
